# Differences in bioprosthetic valve failure after aortic valve replacement in patients with immune mediated inflammatory diseases

**DOI:** 10.1101/2024.07.31.24311322

**Authors:** Christopher Sefton, Davis Leaphart, Benjamin Klein, Garrett Santini, Aditi Patel, M. Elaine Husni, Patrick R. Vargo, Eric E. Roselli, Lars Svensson, Amar Krishnaswamy, Samir Kapadia, Venu Menon, Umesh Khot, Heba Wassif

**Author notes:** **Corresponding Author** Heba Wassif, MD, MPH, Department of Cardiology, Heart Vascular Thoracic Institute, Cleveland Clinic Foundation, 9500 Euclid Ave, J2, Cleveland, OH 44195.

## Abstract

**Introduction:** Immune-mediated inflammatory disease (IMID) is a subset of autoimmune diseases including systemic lupus erythematosus, rheumatoid arthritis, and psoriasis that is emerging as a risk factor for many cardiovascular diseases including valvular disease.

**Objectives:** To determine whether IMID is associated with frequent and early development of bioprosthetic valve failure (BVF) after surgical aortic valve replacement (SAVR) and transcatheter valve replacement (TAVR).

**Methods:** Serial echocardiograms for patients who underwent SAVR and TAVR at Cleveland Clinic between 2000 and 2022 were assessed for time to development of BVF after procedure. ICD10 codes were used to stratify to those with and without IMID. Kaplan-Meier curve and cox proportional hazard regression analysis were used to assess for differences in development of BVF after TAVR and SAVR.

**Results:** 351 TAVR patients (52 IMID and 299 controls) and 1961 SAVR patients (300 IMID and 1661 controls) were included. BVF after TAVR occurred in 12 (23.1%) IMID and 21 (7.0%) control patients, respectively, yielding an adjusted hazard ratio of 4.02 (1.81 - 8.92). Time to 50% of patients developing BVF was earlier among IMID, occurring at 6.6 years IMID and not reached in controls (p < 0.001). There were no significant differences in prevalence and time to development of BVF in IMID vs controls after SAVR.

**Conclusion:** After TAVR, BVF occurred earlier and more frequently in patients with IMID than controls. This risk should be included during shared decision making among IMID patients considered for TAVR, and may warrant more frequent monitoring post procedure. These differences in BVF were not seen after SAVR.

## INTRODUCTION

Immune mediated inflammatory diseases (IMID), including systemic lupus erythematosus (SLE), rheumatoid arthritis (RA), and psoriasis among others^1^, are emerging as significant risk factors for development of early and accelerated cardiovascular disease^2,3,4,5^. Notable examples include increased incidence of cardiovascular disease and cardiovascular deaths in SLE patients^6^, and RA with similar risk for developing coronary artery disease (CAD) as diabetes mellitus^7^. Furthermore, a recent population-based study found increased incidence of nearly every cardiovascular disease, including CAD, atrial fibrillation, heart failure, and valvular disease, in patients with autoimmune disease^8^.

The relationship between valvular disease and IMID has not been fully explored. Recent study suggests there is an increased risk for development of valvular disease in patients with IMID^8,9,10^. One large population-based study reported non-inferior outcomes in acute mortality among IMID patients after surgical aortic valve replacement (SAVR) or transcatheter aortic valve replacement (TAVR) as compared to those without concomitant IMID^11^. However, the role of auto-immune inflammation on the durability of implanted valves remains unclear. Whether patients with IMID are at risk of early prosthetic valve dysfunction or failure compared to patients without IMID is unknown. We sought to explore this question using longitudinal echocardiographic data to assess differences in development of bioprosthetic valve failure (BVF) after surgical and transcatheter aortic valve replacements between patients with and without IMID.

## METHODS

Patients who underwent TAVR or SAVR at Cleveland Clinic between 2000 and 2022 were considered for inclusion. Patients were only included if they had at least 2 echocardiograms in the Cleveland Clinic system available for review after TAVR or SAVR. Patients were included in the IMID group if they had an ICD10 code for autoimmune disease in their chart. These diagnoses included rheumatoid arthritis, psoriasis, vasculitis, polymyalgia rheumatica, Sjogren’s, systemic lupus erythematosus, mixed connective tissue disease, and ankylosing spondylitis (supplemental table 1). Patients without ICD10 codes for IMID were considered controls. Patient demographics, comorbidities and medications were extracted from the medical record.

Serial echocardiograms performed after aortic valve replacement (AVR) were examined for evidence of bioprosthetic valve failure (BVF) after TAVR or SAVR. Patients were determined to have BVF if they developed one of the following: 1) increase in mean transvalvular gradient of greater than or equal to 20 mmHg resulting in a mean gradient greater than or equal to 30 mmHg when compared with gradients 1 to 3 months post procedure; 2) new occurrence of severe aortic regurgitation not from paravalvular leak; or 3) repeat aortic valve intervention. These criteria were derived from previously published criteria from the VARC3 committee^12^. Post-TAVR echocardiograms with concern for aortic regurgitation were manually reviewed to exclude regurgitation due to paravalvular leak. Echocardiograms performed within the Cleveland Clinic enterprise undergo regular quality assurance to ensure consistency in interpretation. Total follow up time for each patient was determined using the most recent echocardiogram performed after aortic valve replacement.

Differences in time to development of BVF after TAVR and SAVR between patients with IMID and without IMID were examined. Patients were included if they developed evidence of BVF at any point after 30 days post-procedure or if they had echocardiogram follow up of at least three years post-procedure. Time to development of BVF was assessed in a univariate capacity with Kaplan-Meier curves using the *survfit* package in R Version 2023.12.0+369 (2023.12.0) as has been previously described^13^. Cox proportional hazards regression was used for multivariate analysis to account for differences in age, sex, and medical comorbidities between the groups as been previously described^13^. Separate analysis was done in the SAVR and TAVR populations. A Kaplan-Meier curve was generated with both TAVR and SAVR patients to compare overall rates of BVF between groups.

## RESULTS

### Bioprosthetic Valve Failure after TAVR

A total of 354 IMID patients and 1202 controls who underwent native valve TAVR at Cleveland Clinic between 2000 and 2022 who had greater than 2 echocardiograms were considered for inclusion in the study. 1,203 (302 IMID and 903 controls) were excluded due to insufficient duration of follow echocardiograms up post TAVR (less than 3 years). 351 TAVR patients (52 IMID and 299 controls) were included in the final analysis (Figure 1A). A higher percentage of IMID patients were excluded compared with controls (302/354 (85.3% excluded) vs 903/1203 (75.1% excluded); p < 0.001). Mean total follow up times were similar for IMID and controls at 4.1±1.4 years and 4.3±1.3 years (p = 0.354), respectively. Medical comorbidities between the IMID and controls were similar, including similar rates of hypertension, hyperlipidemia, chronic kidney disease, coronary artery disease, and atrial fibrillation. Chronic obstructive pulmonary disease and type 2 diabetes were more common in IMID group than controls. Additionally, there were more women in the IMID group (33 (63.5%) vs 114 (38.1%); p <0.001) and the IMID group was younger relative to the controls (78.1±9.4 years vs 80.1±8.7 years; p = 0.017).

**Figure 1A.**
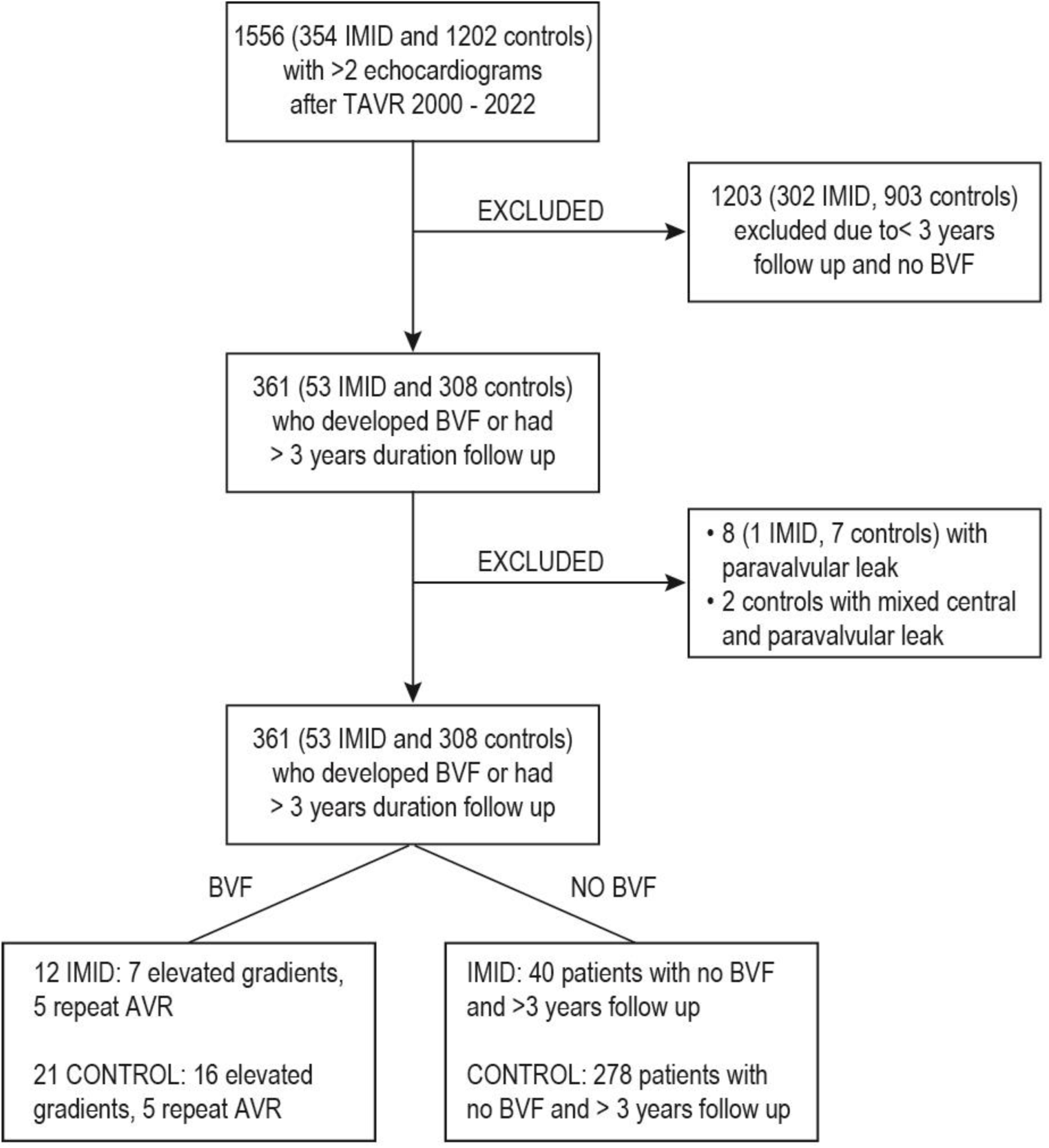
Flow chart delineating inclusion exclusion process for TAVR patients for assessment of BVF. TAVR=transcatheter aortic valve replacement; IMID=immune mediated inflammatory disease; BVF=bioprosthetic valve failure; AVR=aortic valve replacement

**Figure 1B.**
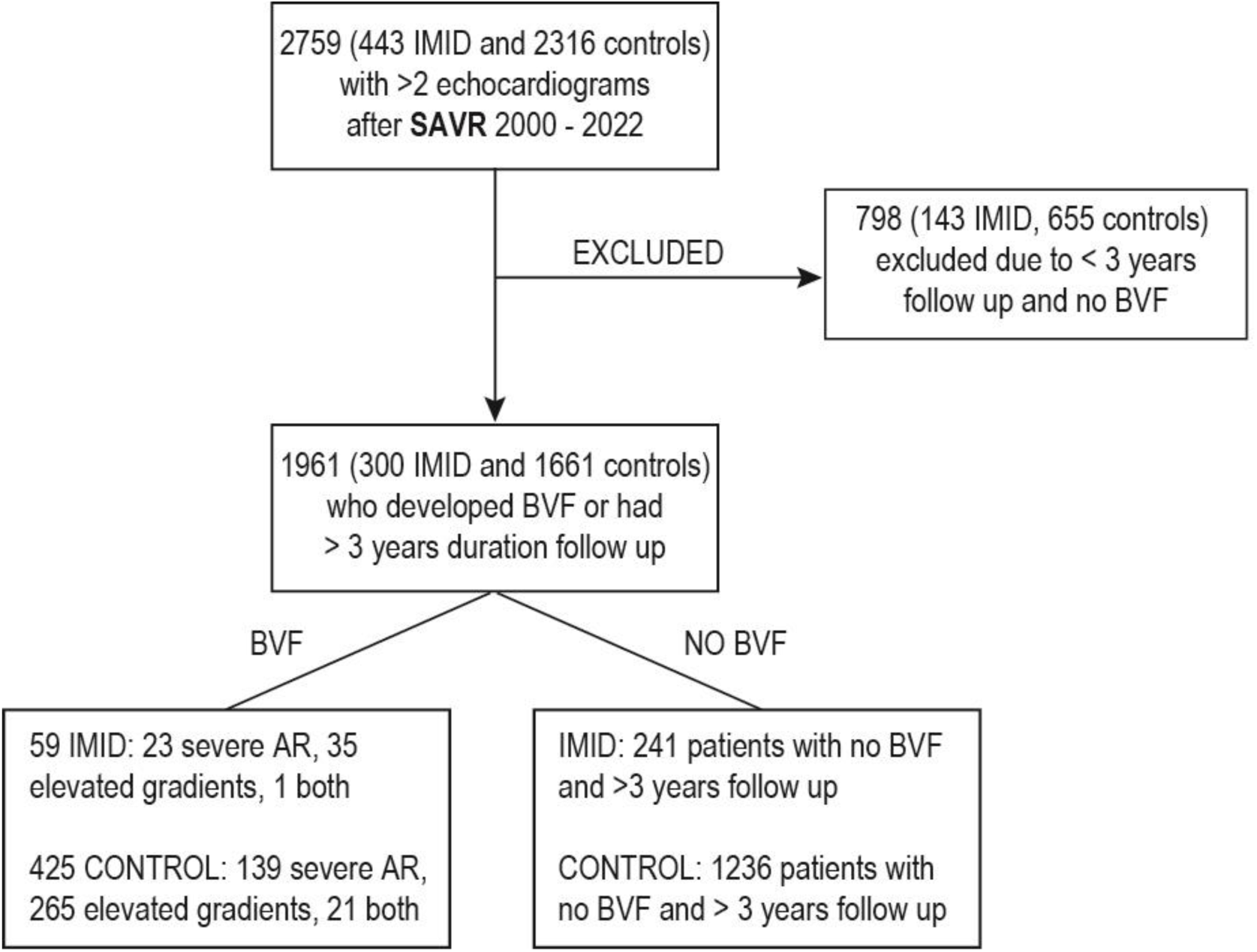
Flow chart delineating inclusion exclusion process SAVR patients for assessment of BVF. SAVR=surgical aortic valve replacement; BVF=bioprosthetic valve failure; IMID=immune mediated inflammatory disease; AR=aortic regurgitation

Medication use and individual autoimmune disease prevalence for IMID patients who underwent TAVR are displayed in table 1 and 2, respectively. Diagnoses included RA (15, 28.8%), psoriasis (12, 23.1%), vasculitis (7, 13.5%), polymyalgia rheumatica (PMR)(7, 120213.5%), mixed connective tissue disease (MCTD) (3, 5.8%), SLE (3, 5.8%), Sjogren’s disease (4, 7.7%), and ankylosing spondylitis (1, 1.9%) (Table 2). Due to low sample size, there was insufficient power to detect differences in BVF for individual IMID diagnoses. Rates of disease-modifying antirheumatic drugs (DMARDs) were predictably higher among IMID patients (Table 1). There were 8 (15.4%) patients on methotrexate, 14 (26.9%) hydroxychloroquine, 5 (9.6%) rituximab, 3 (5.8%) leflunomide, and 4 (7.7%) anti-TNF agents (Table 1).

**Table 1.** Demographics, comorbidities, and medication usage of patients with and without IMID undergoing TAVR and SAVR. P values for TAVR and SAVR are comparing characteristics of IMID versus controls. The third column is comparing the IMID groups between TAVR and SAVR cohorts TAVR=transcatheter aortic valve replacement; SAVR=Surgical aortic valve replacement; IMID=Immune mediated inflammatory disease; SD=standard deviation; COPD=chronic obstructive pulmonary disease; CKD=chronic kidney disease; CAD=coronary artery disease; MI=myocardial infarction; PCI=percutaneous coronary intervention; CABG=coronary artery bypass graft; ARNI=angiotensin receptor/neprilysin inhibitor; SGLT2=sodium glucose transporter inhibitor

|  | TAVR |  |  | SAVR |  |  | IMID in TAVR vs SAVR |  |  |
| --- | --- | --- | --- | --- | --- | --- | --- | --- | --- |
|  | IMID<br>(n= 52) | Control<br>(n = 299) | P-value | IMID<br>(n = 300) | Control<br>(n = 1661) | P-value | IMID TAVR<br>(n = 52) | IMID SAVR<br>(n = 300) | P-value |
| Age, mean (SD) | 78.1 (9.4) | 80.1 (8.7) | **0.017** | 69.3 (12.1) | 66.9 (12.4) | **0.003** | 78.1 (9.4) | 69.3 (12.1) | **<0.001** |
| Female, n (%) | 33 (63.5%) | 114 (38.1%) | **<0.001** | 117 (39.0%) | 451 (27.2%) | **<0.001** | 33 (63.5%) | 117 (39.0%) | **<0.001** |
| Race, n (%) |  |  |  |  |  |  |  |  |  |
| White | 49 (94.2%) | 275 (91.9%) | 0.678 | 281 (93.7%) | 1510 (90.9%) | 0.284 | 49 (94.2%) | 281 (93.7%) | 0.391 |
| Black | 3 (5.8%) | 20 (6.7%) |  | 11 (3.7%) | 93 (5.6%) |  | 3 (5.8%) | 11 (3.7%) |  |
| Other | 0 | 4 (1.3%) |  | 8 (2.7%) | 58 (3.5%) |  | 0 | 8 (2.7%) |  |
| Hypertension, n (%) | 51 (98.1%) | 289 (96.7%) | 0.587 | 283 (94.3%) | 1547 (93.1%) | 0.445 | 51 (98.1%) | 283 (94.3%) | 0.258 |
| Hyperlipidemia, n (%) | 51 (98.1%) | 277 (92.7%) | 0.144 | 269 (89.7%) | 1442 (86.8%) | 0.173 | 51 (98.1%) | 269 (89.7%) | 0.052 |
| COPD, n (%) | 27 (51.9%) | 112 (37.5%) | 0.049 | 134 (44.7%) | 486 (29.3%) | **< 0.001** | 27 (51.9%) | 134 (44.7%) | 0.332 |
| Type 2 Diabetes, n (%) | 27 (51.9%) | 111 (37.1%) | **0.043** | 134 (44.7%) | 598 (36.0%) | **0.004** | 27 (51.9%) | 134 (44.7%) | 0.332 |
| CKD, n (%) | 26 (50.0%) | 151 (50.5%) | 0.947 | 124 (41.3%) | 589 (35.5%) | 0.052 | 26 (50.0%) | 124 (41.3%) | 0.243 |
| CAD, n (%) | 26 (50.0%) | 161 (53.8%) | 0.608 | 154 (51.3%) | 817 (49.2%) | 0.494 | 26 (50.0%) | 154 (51.3%) | 0.859 |
| MI | 18 (34.6%) | 83 (27.8%) | 0.313 | 63 (21.0%) | 300 (18.1%) | 0.228 | 18 (34.6%) | 63 (21.0%) | **0.031** |
| PCI | 8 (15.4%) | 79 (26.4%) | 0.089 | 32 (10.7%) | 181 (10.9%) | 0.906 | 8 (15.4%) | 32 (10.7%) | 0.322 |
| CABG | 11 (21.2%) | 99 (33.1%) | 0.086 | 122 (40.7%) | 677 (40.8%) | 0.976 | 11 (21.2%) | 122 (40.7%) | 0.007 |
| Atrial Fibrillation, n (%) | 32 (61.5%) | 173 (57.9%) | 0.619 | 194 (64.7%) | 1039 (62.6%) | 0.485 | 32 (61.5%) | 194 (64.7%) | 0.664 |
| Tobacco Use, n (%) | 27 (51.9%) | 185 (61.9%) | 0.176 | 177 (59.0%) | 1002 (60.3%) | 0.666 | 27 (51.9%) | 177 (59.0%) | 0.339 |
| Medications, n (%) |  |  |  |  |  |  |  |  |  |
| ARNI | 2 (3.8%) | 27 (9.0%) | 0.211 | 6 (2.0%) | 75 (4.5%) | **0.044** | 2 (3.8%) | 6 (2.0%) | 0.411 |
| SGLT2 | 9 (17.3%) | 27 (9.0%) | 0.069 | 23 (7.7%) | 119 (7.2%) | 0.757 | 9 (17.3%) | 23 (7.7%) | **0.026** |
| Digoxin | 13 (25.0%) | 40 (13.4%) | **0.031** | 57 (19.0%) | 268 (16.1%) | 0.219 | 13 (25.0%) | 57 (19.0%) | 0.317 |
| Methotrexate | 8 (15.4%) | 1 (0.3%) | **<0.001** | 52 (17.3%) | 7 (0.4%) | **<0.001** | 8 (15.4%) | 52 (17.3%) | 0.731 |
| Hydroxychloroquine | 14 (26.9%) | 0 | **<0.001** | 42 (14.0%) | 10 (0.6%) | **<0.001** | 14 (26.9%) | 42 (14.0%) | **0.019** |
| Rituximab | 5 (9.6%) | 1 (0.3%) | **<0.001** | 9 (3.0%) | 0 | **<0.001** | 5 (9.6%) | 9 (3.0%) | **0.024** |
| Leflunimide | 3 (5.8%) | 0 | **<0.001** | 14 (4.7%) | 0 | **<0.001** | 3 (5.8%) | 14 (4.7%) | 0.732 |
| Anti-TNF | 4 (7.7%) | 0 | **<0.001** | 26 (8.7%) | 9 (0.5%) | **<0.001** | 4 (7.7%) | 26 (8.7%) | 0.816 |

**Table 2.** Prevalence of individual autoimmune diagnoses in TAVR and SAVR cohort. P value determined in assessing the difference in distribution of each individual diagnosis between the TAVR and SAVR cohort SLE=systemic lupus erythematous; PMR=polymyalgia rheumatica; TAVR=Transcatheter aortic valve replacement; SAVR=surgical aortic valve replacement

| Disease, n (%) | TAVR (n =52) | SAVR (n=300) | P - value |
| --- | --- | --- | --- |
| Rheumatoid Arthritis | 15 (28.8%) | 98 (32.7%) | 0.586 |
| SLE | 3 (5.8%) | 17 (5.7%) | 0.976 |
| Sjogren's | 4 (7.7%) | 15 (5.0%) | 0.428 |
| PMR | 7 (13.5%) | 40 (13.3%) | 0.968 |
| Vasculitis | 7 (13.5%) | 41 (13.6%) | 0.728 |
| Mixed Connective<br>Tissue Disease | 3 (5.8%) | 17 (5.7%) | 0.976 |
| Psoriasis | 12 (23.1%) | 65 (21.7%) | 0.821 |
| Ankylosing Spondylitis | 1 (1.9%) | 7 (2.3%) | 0.855 |

Edwards LifeSciences was the most common manufacturer represented among the cohort. Medtronic, Abbot, Boston Scientific, and Direct Flow represented a small minority of valves present. Edwards Lifesciences valves were less prevalent among IMID compared to controls (43 (82.7%) vs 276 (92.3%); p = 0.026) Mean valve size was smaller among IMID patients relative to control patients (24.7±2.6 vs 25.7±2.5; p = 0.005) (Table 4).

The development of BVF after TAVR was more frequent in patients with IMID when compared with controls. BVF occurred in 12 (23.1%) of IMID patients compared with 21 (7.0%) of controls. Among IMID patients, 5 met BVF criteria due to repeat aortic valve replacement and 7 developed BVF due to severely increased gradients. Indication for repeat aortic valve replacement was infective endocarditis (IE) in 4 patients, and severe prosthetic valve aortic regurgitation in 1 patient. One IMID patient developed aortic regurgitation, but upon manual review this was paravalvular leak and was excluded. Among controls, 5 met BVF criteria due to repeat aortic valve intervention and 16 developed BVF due to increased gradients. Indication for repeat aortic valve replacement was IE in 2 patients and severe prosthetic valve stenosis in 3 patients. There were 9 patients with evidence of aortic regurgitation, but upon manual review 7 had a component of paravalvular leak and were excluded. For consistency, the remaining 2 were excluded in order to have consistent comparisons between IMID and controls within the TAVR comparison.

Development of BVF also occurred significantly earlier in IMID patients compared with controls. Figure 2 shows a Kaplan Meier curve demonstrating earlier failure rates in IMID, with estimated time to 50% of IMID developing BVF to be 6.6 years, while this time could not be calculated in controls due to low prevalence of failure (p < 0.001) (Figure 2). Differences in development of BVF persisted after multivariate analysis with cox proportional hazard regression. After adjustment for differences in age, sex, and comorbidities between IMID and control group, the adjusted hazard ratio for developing BVF in IMID compared with control was 4.02 (CI: 1.81 – 8.92) (p < 0.001) (Table 3). Development of BVF was also independently associated with increased age and nonwhite race (Table 3).

**Figure 2.**
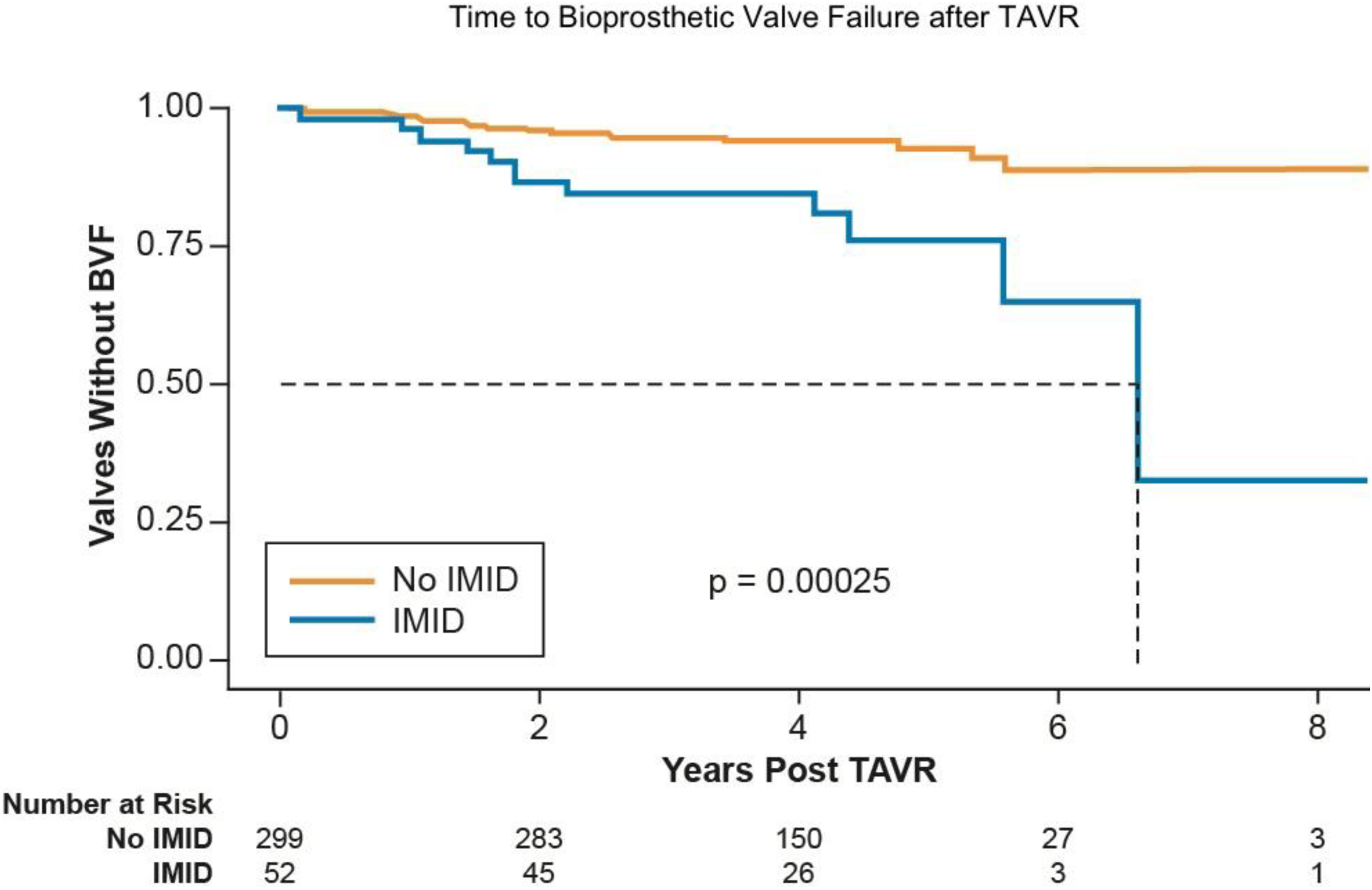
Kaplan Meier curve assessing development of BVF after TAVR in patients with and without IMID. IMID=immune mediated inflammatory disease; TAVR=transcatheter aortic valve replacement; BVF=bioprosthetic valve failure

**Table 3.** Risk factors for developing BVF after TAVR and SAVR. Adjusted hazards ratios generated using cox proportional hazards regression in multivariate fashion IMID=immune mediated inflammatory disease; TAVR=transcatheter aortic valve replacement; SAVR=surgical aortic valve replacement; AHR=adjusted hazard ratio; COPD=chronic obstructive pulmonary disease

|  | TAVR |  | SAVR |  |
| --- | --- | --- | --- | --- |
|  | AHR (95% CI) | P-value | AHR (95% CI) | p-value |
| IMID | **4.02** (1.81 - 8.92) | **6.15E-04** | 0.90 (0.68-1.19) | 0.477 |
| Male Sex | 0.97 (0.45-2.07) | 0.932 | **0.75** (0.59-0.88) | **0.005** |
| White Race | **0.32** (0.12 – 0.91) | **0.032** | 0.74 (0.53-0.97) | 0.061 |
| Age | **0.97** (0.94 – 0.99) | **0.042** | **0.98** (0.97-0.99) | **7.48E-06** |
| Hypertension | 1.36 (0.18 – 10.43) | 0.767 | 1.26 (0.79-1.62) | 0.224 |
| Hyperlipidemia | 0.73 (0.22 – 2.40) | 0.601 | **0.72** (0.54-0.91) | **0.012** |
| COPD | 0.56 (0.25 – 1.24) | 0.155 | 1.11 (0.91-1.34) | 0.294 |
| Type 2 Diabetes | 0.81 (0.37 – 1.81) | 0.597 | 1.11 (0.89-1.32) | 0.301 |
| Chronic Kidney Disease | 1.16 (0.55 – 2.48) | 0.687 | 1.10 (0.90-1.34) | 0.384 |
| Atrial Fibrillation | 0.89 (0.43 – 1.86) | 0.768 | 1.12 (0.91-1.33) | 0.329 |
| Coronary Artery Disease | 0.65 (0.29 – 1.41) | 0.277 | **1.30** (1.07-1.58) | **0.009** |

### Bioprosthetic valve failure after SAVR

There were 443 patients with IMID and 2316 controls who underwent native valve SAVR between 2000 and 2022 with at least 2 consecutive echocardiograms post procedure in the Cleveland Clinic system. Of these, 798 (143 IMID and 655 controls) were excluded due to insufficient follow up time of less than 3 years. A total of 1961 (300 IMID and 1661 controls) were included in the analysis after exclusions. A higher percentage of IMID were excluded relative to controls (143/443 (32.3% excluded) vs 655/2316 (28.3% excluded); p = 0.089). Comorbidities such as HTN, HLD, HLD, CAD, and atrial fibrillation were similar between groups. When comparing IMID to controls, there was more type 2 diabetes (134 (44.7%) vs 598 (36.0%); p = 0.004), COPD (131 (43.7%) vs 486 (29.3%); p < 0.001), and women (117 (39.0%) vs 451 (27.2%); p < 0.001). The IMID group was also older, with mean age 69.3±12.1 years vs 66.9±12.4 years (p = 0.003). Similar to the TAVR analysis, rates of disease-modifying antirheumatic drugs (DMARDs) was predictably higher among IMID patients (Table 1).

Medication use and individual autoimmune disease prevalence for IMID patients who underwent SAVR can be seen in table 1 and 2, respectively. Diagnoses included RA 98, 32.7%), psoriasis (65, 21.7%), vasculitis (41, 13.7%), PMR (40, 13.3%), MCTD (17, 5.7%), SLE (17, 5.7%), Sjogren’s (15, 5.0%), and ankylosing spondylitis (7, 2.3%) (Table 2). There were 52 (17.3%) patients on methotrexate, 42 (14.0%) hydroxychloroquine, 9 (3.0%) rituximab, 14 (3.0%) leflunomide, 26 (8.7%) anti-TNF agents (Table 1).

Bioprosthetic, homograft, and mechanical valve replacements were present in both groups. Bioprosthetic replacement was most common, and homograft and mechanical replacements had similar prevalence (Table 5). Prevalence of each replacement type was similar in IMID compared with control, with bioprosthetic being slightly more prevalent among IMID compared with controls (p = 0.074)(Table 5). Manufacturers of each valve type were collected for each group. Among bioprosthetic valves, Edwards Lifesciences and St. Jude Trifecta were the most common manufacturers. Several additional manufacturers made up a small minority of the remainder of the valves, including Cryolife, Medtronic, Perceval, and Sorin. Overall prevalence of each manufacturer for IMID and control valves were similar among bioprosthetic valves (p = 0.812)(Table 4). Among mechanical valve replacements, St Jude, Carbomedics, and On-X were present with distribution of individual manufacturers being similar when IMID group is compared to control (p = 0.942)(Table 5). Mean valve size was smaller in IMID compared with controls (23.0±2.3 vs 23.3±2.4; p = 0.0299).

**Table 4.** Characteristics of aortic valves used in transcatheter aortic valve replacements among patients with and without IMID. P value assess distribution between Edwards Lifesciences, Medtronic, and Other manufacturers. Valve types in italics below Edwards Lifesciences and Other are subsets of those categories included in the total IMID=Immune mediated inflammatory disease; SD=Standard deviation

|  | IMID (n = 52) | Control (n = 299) | P value |
| --- | --- | --- | --- |
| Valve Size (mm),<br>mean (SD) | 24.7 (2.6) | 25.7 (2.5) | 0.005 |
| **Valve Manufacturer** |  |  | 0.026 |
| **Edwards Lifesciences** | 43 (82.7%) | 276 (92.3%) |  |
| *Sapien 3* | 41 (76.9%) | 271 (90.6%) |  |
| *Sapien XT* | 2 (3.8%) | 3 (1.0%) |  |
| *Sapien Ultra* | 0 | 2 (0.6%) |  |
| **Other** | 9 (17.3%) | 23 (7.7%) |  |
| *Medtronic* | 7 (13.4%) | 12 (4.0%) |  |
| *Boston Scientific* | 1 (1.9%) | 4 (1.3%) |  |
| *Direct Flow* | 1 (1.9%) | 6 (2.0%) |  |
| *Abbot* | 0 | 1 (0.3%) |  |

**Table 5.**
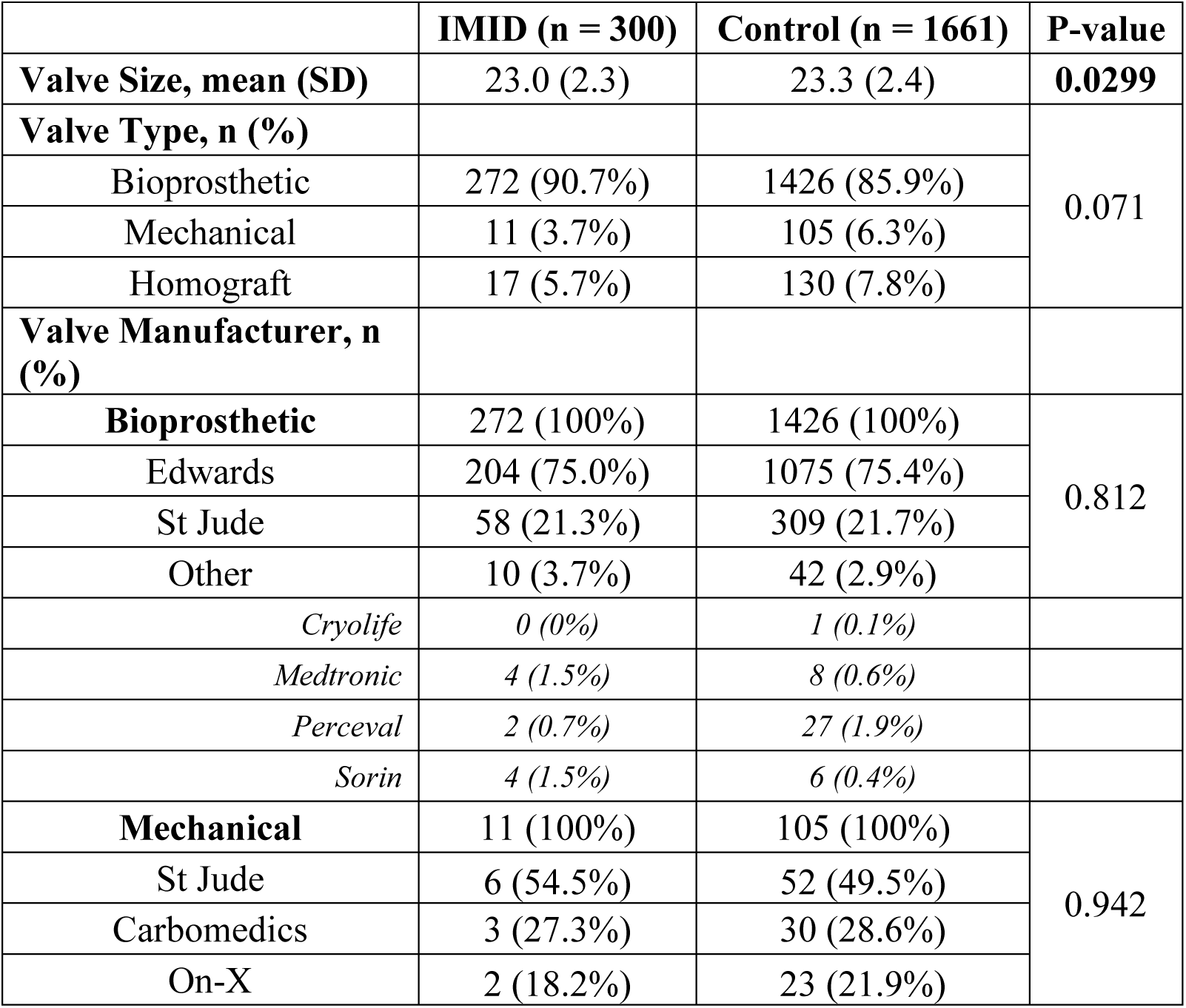
Characteristics of aortic valves used in surgical aortic valve replacements among patients with and without IMID. P values compare total distribution of BP, mechanical, and homograft valves as well as the distribution of types of valves within the bioprosthetic and mechanical subsets IMID=immune mediate inflammatory disease; SD=standard deviation;

The mean follow up time in patients who underwent SAVR included for study was 8.9±4.6 years and 9.5±5.3 years for IMID and controls, respectively (p = 0.056). There was no significant difference in the development BVF in the IMID group compared with control. BVF occurred in 59 (19.7%) of IMID patients and in 425 (25.6%) of control patients. Among BVF in IMID patients, 23 had severe aortic regurgitation, 35 had elevated gradients, and one had evidence of both at first detection of BVF. Among controls, 139 had severe aortic regurgitation, 265 had elevated gradients, and 21 had evidence of both at first detection of BVF. Multivariable analysis was performed with cox proportional hazard regression. After adjustment for differences in age, sex, and comorbidities between IMID and control group, the adjusted hazard ratio for developing BVF in IMID compared with control was 0.90 (CI: 0.68 – 1.19) (p = 0.417) (Table 3). Additionally, female sex, increased age, and CAD were associated with increased risk of developing BVF independent of IMID status (Table 3). Estimated time to development of BVF after SAVR was also not significantly different, occurring at 16.2 years and 16.0 years in the IMID and control groups, respectively (p = 0.47). Kaplan Meier curve demonstrating development of BVF after SAVR over time in both groups is displayed in Figure 3.

**Figure 3.**
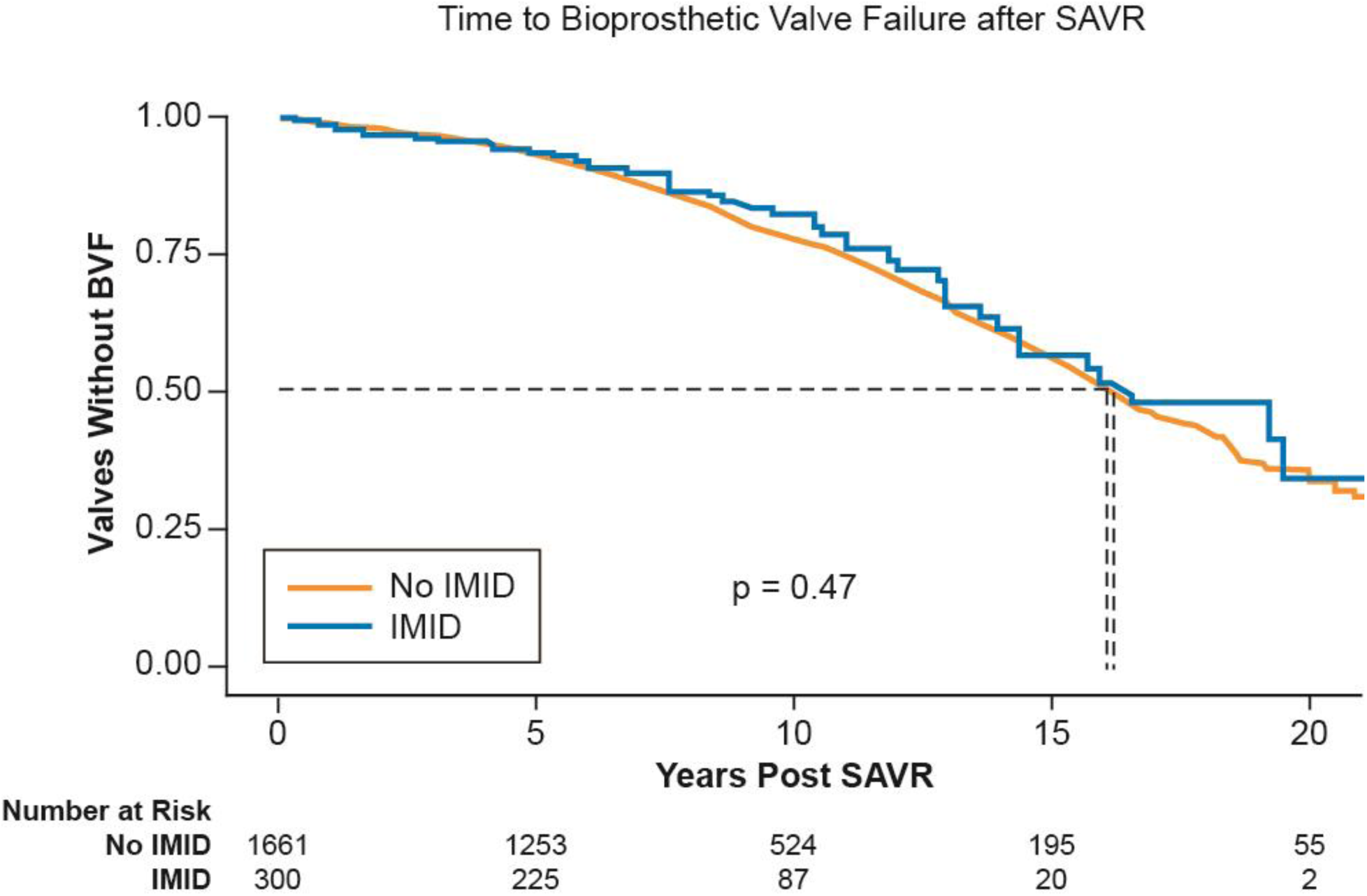
Kaplan Meier curve assessing development of BVF after SAVR in patients with and without IMID. IMID=immune mediated inflammatory disease; SAVR=surgical aortic valve replacement; BVF=bioprosthetic valve failure

RA was the most common IMID diagnosis in the SAVR cohort, and a separate analysis was performed to assess development of BVF after SAVR in RA patients only compared to controls. Within the IMID SAVR cohort, there were 98 patients with RA available for study. There was no difference in time to development of BVF after SAVR in in patients with RA compared to patients without RA (Figure 4).

**Figure 4.**
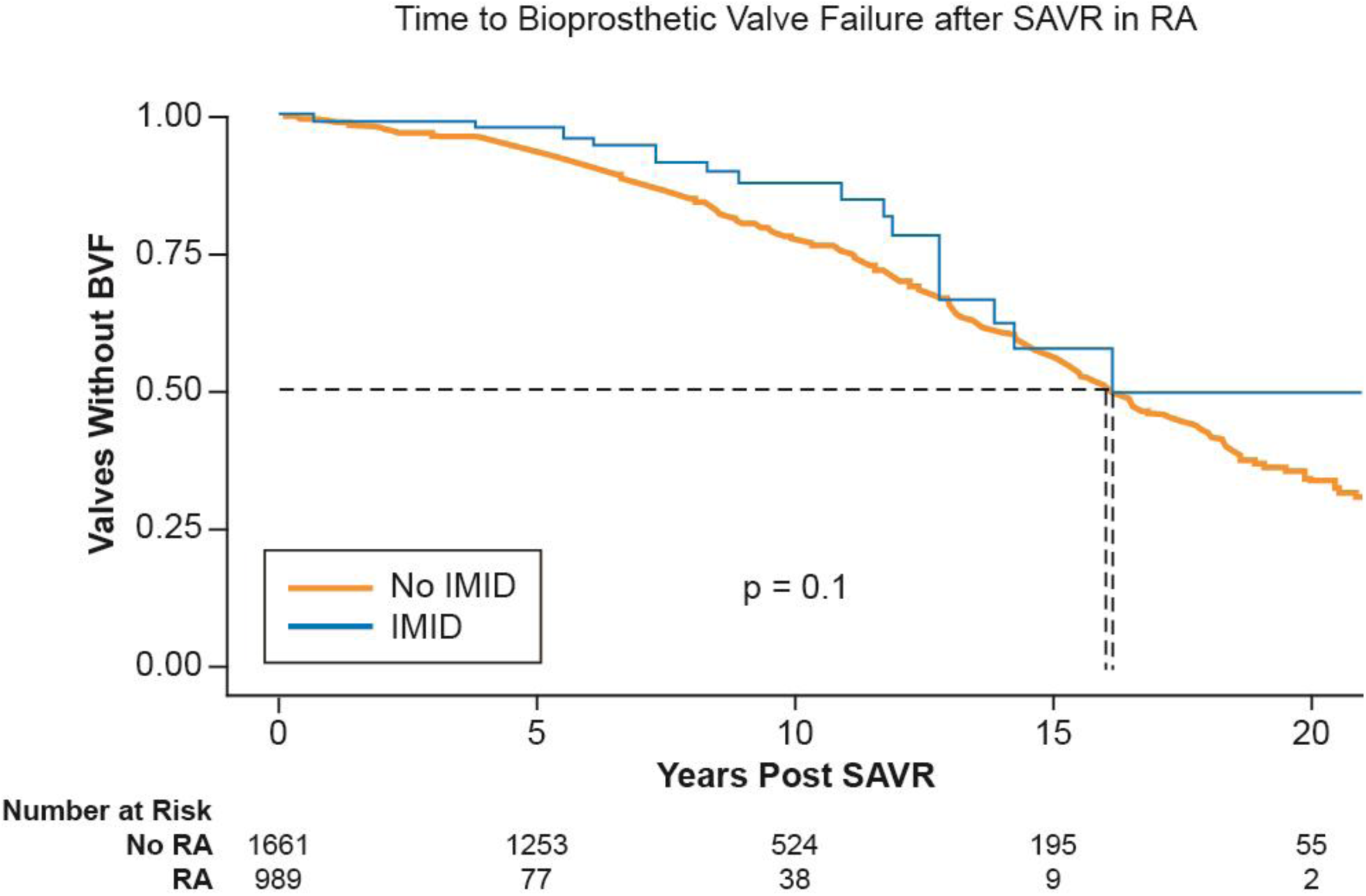
Kaplan Meier curve assessing development of BVF after SAVR in patients with and without Rheumatoid Arthritis. RA=rheumatoid arthritis; SAVR=surgical aortic valve replacement; BVF=bioprosthetic valve failure

### Comparing IMID population in TAVR and SAVR

Demographic, medical comorbidities, medication use, and individual diagnoses within patients with IMID who underwent TAVR and SAVR were compared (Table 1)(Table 2). Notably, when compared with SAVR IMID patients, TAVR IMID patients were significantly older (mean age 78.1±9.4 vs 69.3±12.1 years; p < 0.001) and contained higher proportion of females (33 (63.5%) vs 117 (39.0%); p < 0.001), though the trend of more females in the IMID group was the same in both TAVR and SAVR. Comorbidities were similar between groups. Prevalence of CAD was similar between groups, but rates of prior myocardial infarction were higher among TAVR patients (18 (34.6%) vs 63 (21.0%); p = 0.031). There was significantly higher prevalence of hydroxychloroquine usage among TAVR patients compared with SAVR patients (14 (26.9%) vs 42 (14.0%); p = 0.019) as well as rituximab (5 (9.6%) vs 9 (3.0%); p = 0.024). Usage of other DMARDs was similar. Rates of individual autoimmune disease diagnoses were similar between groups (Table 2).

Development of BVF occurred earlier in the IMID group in the TAVR cohort but not in the SAVR cohort as discussed above. TAVR cohort members only had bioprosthetic valves, while SAVR cohort members were distributed between bioprosthetic (majority), mechanical, and homografts. In order to assess for confounding of the mechanical and homograft valves in the SAVR cohort, a separate analysis was performed using only the patients who had bioprosthetic valves in the SAVR cohort and comparing this with the TAVR cohort. This generated a Kaplan Meier curve (Figure 5) with both SAVR (bioprosthetic valves only) and TAVR cohorts together. TAVR patients with IMID developed BVF significantly earlier than the rest of the groups. TAVR patients without IMID appear to develop BVF at a similar rate to the SAVR cohort (Figure 5).

**Figure 5.**
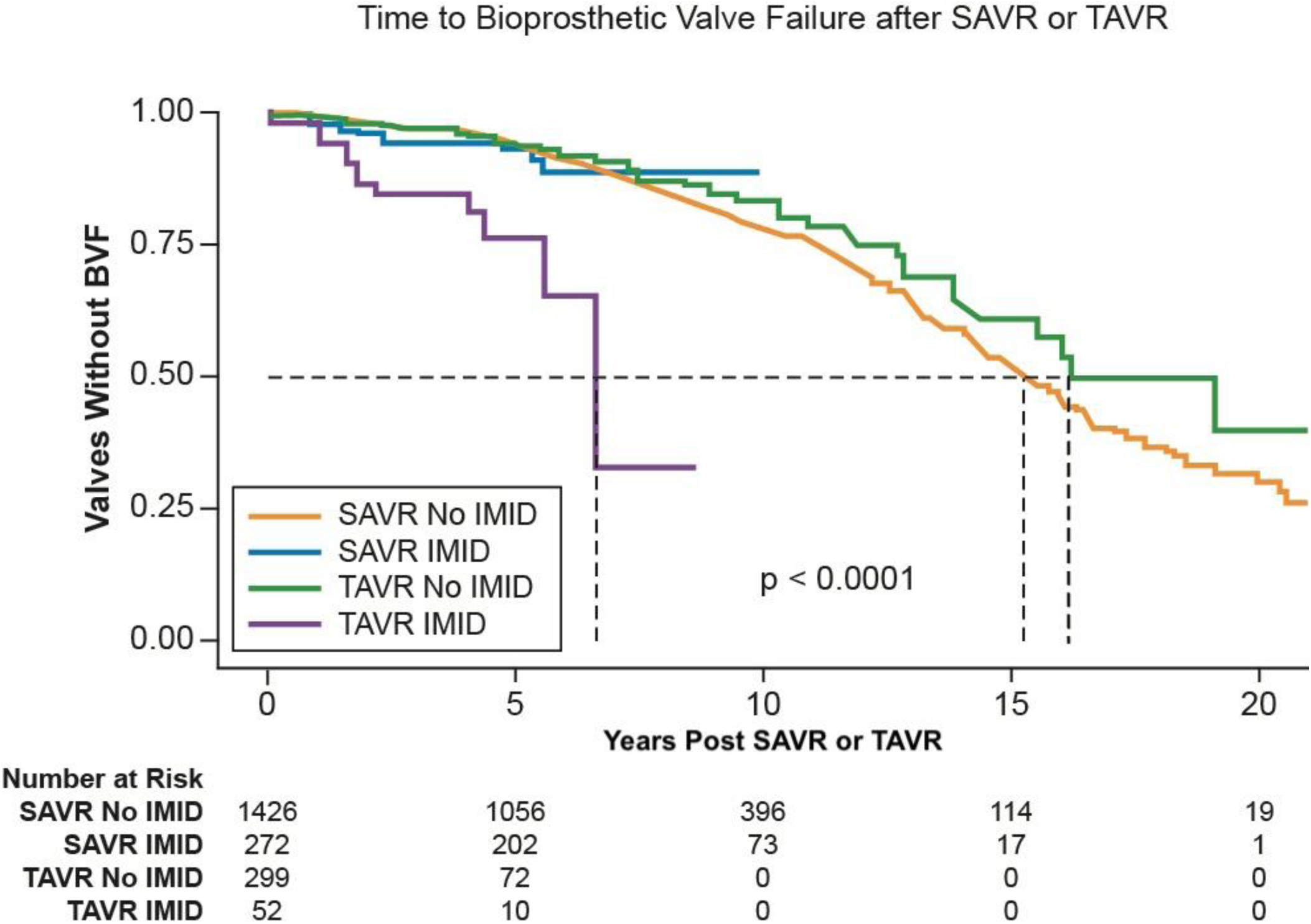
Kaplan Meier curve assessing development of BVF after SAVR or TAVR in patients with and without IMID. SAVR=surgical aortic valve replacement; TAVR=Transcatheter aortic valve replacement; IMID=immune mediate inflammatory disease; BVF=bioprosthetic valve failure; AVR=Aortic valve replacement

## DISCUSSION

To our knowledge, this is the first study to examine bioprosthetic valve function after TAVR and SAVR in patients with IMID. Our results support that patients with IMID who undergo TAVR develop BVF significantly earlier and more frequently relative to patients without IMID. These differences in BVF among IMID patients were not seen after SAVR, but comparisons between the studied TAVR and SAVR populations are limited by the retrospective, observational nature of the study. Prior population-based studies examining outcomes in patients with autoimmune disease after aortic valve replacement have focused on outcomes such as mortality, length of hospital stay, and readmission using broad national databases^11, 14^. They showed similar rates of mortality with the primary conclusion that autoimmune disease should not preclude this patient population from undergoing these procedures. The results of the present study are important, because they suggest IMID patients have different outcomes in prosthetic valve function after TAVR than those without IMID. While IMID should not preclude patients from undergoing these procedures, it may necessitate special consideration in choosing between TAVR or SAVR in these patients. Frequency of follow-up imaging in the IMID patients may also be impacted by these findings.

Notably, the TAVR IMID group was the only group studied with significantly different rates of BVF, and differences in BVF in patients with or without IMID were not observed in the SAVR cohort (Figure 5). The control TAVR group developed BVF at similar rates to all SAVR groups. This is consistent with growing evidence that long term outcomes of TAVR are similar to SAVR^15^, and this consistency lends strength to this study.

Differences in characteristics between the IMID TAVR and SAVR groups were considered in an attempt to explain this difference in outcomes between AVR methods. Rates of individual IMID diagnoses were similar within the groups, as were majority of medical comorbidities, other than SAVR patients being notably younger. Younger age has consistently been shown to be associated with BVF in prior surgical studies^16^. The only rheumatologic difference observed between groups was rate of hydroxychloroquine and rituximab utilization. Prevalence of hydroxychloroquine and rituximab in TAVR IMID patients were roughly double that seen in SAVR IMID patients (Table 1). The role of DMARDs in development of cardiovascular disease in patients with autoimmune disease is an area of interest, with many studies suggesting an overall protective effect with increased exposure^17, 18^. The differences in medication usage may signal differences in disease severity, which could contribute to differences in outcomes between the TAVR and SAVR groups.

Differences in bioprosthetic valve characteristics were also considered as potential confounders in the study, including differences in valve sizes and types between TAVR and SAVR groups. On average, IMID patients had smaller valve size in both TAVR and SAVR compared with controls, likely explained by higher proportion of women in the IMID groups. This trend was similar in both TAVR and SAVR cohorts, however the skew toward women was greater among TAVR. When considered in multivariate analysis with cox regression, male sex appeared protective against BVF in SAVR, but not TAVR. This difference between populations could contribute to the differences in outcomes and larger study with sufficient power for more nuanced matching is warranted in the future.

One hypothesis for the development of BVF after TAVR but not SAVR may be related to the mechanism of valvular disease in IMID and the differences in how the aortic valve procedures are performed. The biologic mechanisms of valvular disease from calcification is a result of inflammation from CD4+ T-cell signaling and pro-inflammatory macrophages^19, 20, 21, 22^. Studies reporting surgical pathology of aortic stenosis in patients with autoimmune disease show a more inflammatory microscopic profile relative to what is seen in patients without inflammatory disease^23, 24^. The excision of the native aortic valve and surrounding tissue during SAVR may be protective against future inflammation and degradation in the prosthetic valve, in contrast to TAVR where the native inflamed tissue remains post procedure. Other potential mechanisms could be related to the hypercoagulable state seen in some IMID. Hypoattenuating leaflet thrombosis, a known complication after SAVR and TAVR with unclear clinical implications^25, 26, 27^, was not assessed in the present study but would warrant investigation in future research.

Furthermore, IE was more prevalent among IMID patients after TAVR compared with controls. Differences in development of IE were not seen after SAVR. There were 4 IMID patients (7.7%) and 2 control patients (0.7%) who developed IE post TAVR. The mechanism for this difference is unclear. It could be similarly related to the proinflammatory state of IMID as described in the previous paragraph, leaving the bioprosthetic valve at increased vulnerability for seeding with transient bacteremia due to increased inflammation. Alternatively, the immunosuppression used in management of IMID could predispose these patients to IE. However, the 4 IMID patients who developed IE were either on MTX monotherapy (2 patients) or HCQ monotherapy (2 patients) rather than more severe immunosuppression such as rituximab or other biologic agents.

The difference in outcomes in the TAVR and SAVR groups has potential significance for IMID patients being considered for AVR. Since FDA approval of TAVR in low-risk surgical patients in 2019, the role of TAVR has been expanded to patients who previously would have been recommended SAVR^28^. Continued positive results from long term, prospective follow up such at the NOTION trial^15^ will likely continue to increase rates of TAVR. Additionally, guidelines recommend “shared decision making” for decision of TAVR vs SAVR in patients age 65 to 80^29^. The results of the present study suggest patients with IMID in this age window being considered for TAVR should be carefully counseled on the potential risk of early BVF. Special considerations may be necessary, such as increased frequency of follow up due to increased risk of early BVF. The potential increased risk for IE in IMID patients should also be considered during shared decision making, and antibiotics for IE prophylaxis should be emphasized in this population. Further study is needed in larger cohorts to validate these findings and make interpopulation assessments between differences in SAVR and TAVR outcomes in patients with IMID.

This study has limitations. Notably, the rate of valve dysfunction reported here appears higher than previously reported^15, 30^. This is likely due to the retrospective nature of the study, which artificially increases prevalence of BVF since significant patient numbers were excluded due to insufficient follow up. This preferentially excludes patients without valve issues, since sicker patients would be expected to have closer follow-up. Secondly, while the study effectively captured a multitude of IMID diagnoses, disease severity is not readily accounted for in our analysis. It is possible that comorbid IMID disease severity in TAVR patients was greater than in SAVR patients, which could explain the differences seen in development of BVF. Thirdly, due to high rates of exclusion for lack of appropriate follow up, our population of IMID patients is small. The control TAVR and SAVR patients are also a small subset of a larger cohort patients who have undergone aortic valve replacement at Cleveland Clinic. This small size increases potential for both type 1 and type 2 error, and repeat study is warranted in a larger, multicenter population. Lastly, while both TAVR and SAVR populations were studied, limited comparisons between the groups can be made due to the retrospective, observational nature of the study. Further large-scale study is needed to draw conclusions when comparing patients with IMID who undergo SAVR and TAVR.

In conclusion, in this single-center, retrospective study, patients with IMID that undergo TAVR develop BVF significantly earlier and more frequently than patients without IMID. Rates of BVF after SAVR were similar between patients with and without IMID. This may have implications for shared decision making in choosing aortic valve interventions for patients with IMID, and warrant increased follow up intervals in IMID patients who undergo TAVR.

## Data Availability

Internal Cleveland Clinic Dataset

## Abbreviations

IMID: immune mediated inflammatory disease
SLE: systemic lupus erythematosus
RA: Rheumatoid arthritis
CAD: Coronary artery disease
SAVR: surgical aortic valve replacement
TAVR: Transcatheter aortic valve replacement
BVF: bioprosthetic valve failure
AVR: Aortic valve replacement
MCTD: mixed connective tissue disease
DMARD: disease modifying antirheumatic drug

## Notes

### Competing Interest Statement

The authors have declared no competing interest.

### Funding Statement

No external funding for report

### Author Declarations

Approved by Cleveland Clinic IRB Study Number 23-095

